# Plasma proteomic signatures of adiposity are associated with cardiovascular risk factors and type 2 diabetes risk in a multi-ethnic Asian population

**DOI:** 10.1101/2023.07.25.23293136

**Authors:** Charlie G.Y. Lim, Bige Ozkan, Yujian Liang, Jiali Yao, Nang Ei Ei Khaing, Mary R. Rooney, Chiadi E. Ndumele, E Shyong Tai, Josef Coresh, Xueling Sim, Rob M. van Dam

**Affiliations:** Saw Swee Hock School of Public Health, National University of Singapore and National University Health System, 12 Science Drive 2, Singapore 117549; Department of Epidemiology and the Welch Center for Prevention, Epidemiology, and Clinical Research, Johns Hopkins Bloomberg School of Public Health, Baltimore, Maryland, USA; Ciccarone Center for the Prevention of Cardiovascular Disease, Division of Cardiology, Johns Hopkins University School of Medicine, Baltimore, Maryland, USA; Department of Medicine, Yong Loo Lin School of Medicine, National University of Singapore, 10 Medical Dr, Singapore 117597; Departments of Exercise and Nutrition Sciences and Epidemiology, Milken Institute School of Public Health, The George Washington University, 950 New Hampshire Ave, NW, Washington, DC 20052

## Abstract

The molecular mechanisms connecting obesity and cardiometabolic diseases are not fully understood. We evaluated the associations between body mass index (BMI), waist circumference (WC), and ∼5,000 plasma proteins in the Singapore Multi-Ethnic Cohort (MEC1). Among 410 BMI-associated and 385 WC-associated proteins, we identified protein signatures of BMI and WC and validated them in an independent dataset across two timepoints and externally in the Atherosclerosis Risk in Communities (ARIC) study. The BMI- and WC-protein signatures were highly correlated with total and visceral body fat, respectively. Furthermore, the protein signatures were significantly associated with cardiometabolic risk factors and were able to differentiate between metabolically healthy and unhealthy obesity. In prospective analyses, the protein signatures were strongly associated with type 2 diabetes risk in MEC1 (odds ratio per SD increment in WC-protein signature = 2.84, 95% CI 2.47 to 3.25) and ARIC (hazard ratio = 1.97, 95% CI 1.87 to 2.07). Pathways related to cell signaling, systemic inflammation, and glucose and fat metabolism were overrepresented in the BMI- and WC-protein signatures. Our protein signatures have potential uses for the monitoring of metabolically unhealthy obesity.

**Article Highlights:**

- We evaluated the associations between ∼5000 plasma proteins and BMI and WC in a multi-ethnic Asian population.
- We identified 410 proteins associated with BMI and 385 proteins associated with WC and derived protein signatures of BMI and WC, which we validated externally in a US cohort.
- Both the BMI- and WC-protein signatures were strongly associated with cardiometabolic risk factors and the risk of type 2 diabetes and were enriched in pathways relating to cell signaling, systemic inflammation, and glucose and fat metabolism.
- Our protein signatures have potential uses for monitoring metabolically unhealthy obesity.

---

Obesity is a metabolic disorder resulting from a complex interplay between genetic, psychosocial, and environmental factors (1). Although obesity is a major risk factor for metabolic diseases such as type 2 diabetes and cardiovascular diseases (CVDs) (2), the biological mechanisms connecting obesity and these metabolic diseases are not fully understood.

Proteins are important biomarkers for identifying biological pathways linking genetic and environmental exposures to disease risk (3). Recent studies in European populations have identified proteins associated with body mass index (BMI) (4–6), highlighting pathways involved in lipid metabolism and inflammations that may contribute to obesity-related diseases (5). However, these studies were based on platforms that covered a limited number of proteins (∼1100-3600 proteins), used BMI as the only measure of adiposity, and included participants of predominantly European ancestry. The latter may be particularly important given the well-recognized differences in the association of BMI with type 2 diabetes in populations of Asian compared to European ancestry (7). Finally, previous studies did not examine whether obesity-related proteins are associated with cardiometabolic health.

We evaluated the associations between ∼5000 plasma proteins, and BMI and waist circumference (WC) in a multi-ethnic Asian population comprising adults of Chinese, Malay, and Indian ethnicity. Using machine learning techniques, we developed protein signatures of BMI and WC and evaluated their associations with total and visceral body fat, CVD risk factors, and the incidence of type 2 diabetes.

## Research Design and Methods

### Study design

An overview of this study is shown in **Figure 1**. Participants in this study were sampled from the Singapore Multi-Ethnic Cohort Phase 1 (MEC1) (8). The MEC1 is a population-based cohort comprising 14,465 male and female adults recruited between 2004 and 2010. Oversampling of ethnic minorities (Malay and Indians) was done to achieve a good representation of three major Asian ethnic groups in Singapore—Chinese, Malay, and Indian. Between 2011 to 2016 (mean follow-up of 6.4 years), 6,112 participants agreed to a follow-up visit. Participants completed a standardized interviewer-administered questionnaire on socio-demographic characteristics, lifestyle, and medical history for both the baseline and follow-up study. Height was measured without shoes on a portable stadiometer, while weight was measured on a digital scale. WC was measured by trained research staff using stretch-resistant tape at the mid-point between the last rib and iliac crest. Blood was drawn for biomarker measurements and biobanking at −80°C.

**Figure 1.**
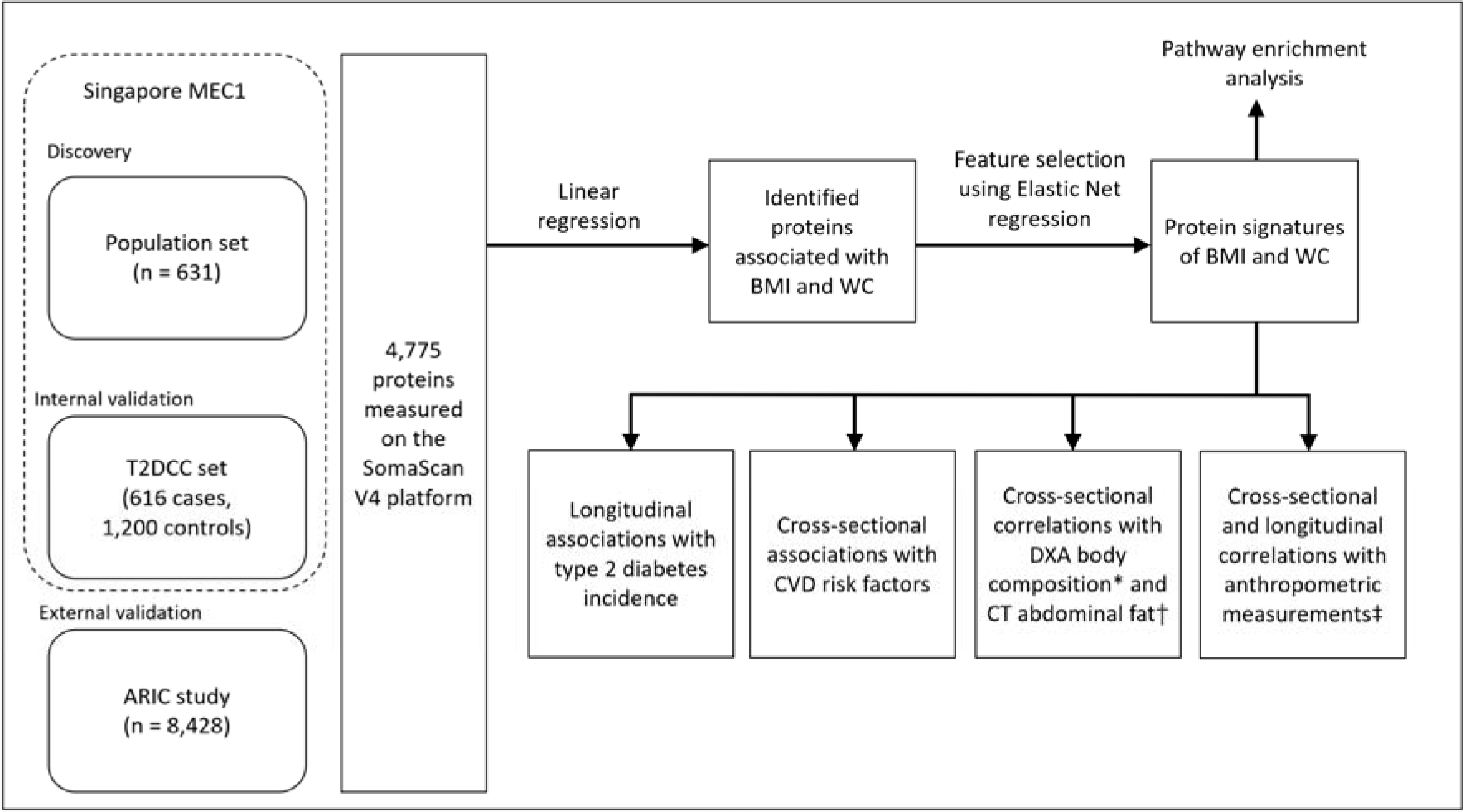
Overview of the study design.* Measured in a subset of Chinese participants in the MEC1 (n = 207) at follow-up. † Measured in a subset of Chinese participants in the MEC1 (n = 151) at follow-up. ‡ Cross-sectional correlations between protein signatures and anthropometric measurements at baseline (n = 1,816) and follow-up (n = 814). Longitudinal correlations between the change in protein signatures and change in anthropometric measurements in participants with data across two timepoints (n = 814). Abbreviations: ARIC, Atherosclerosis Risk in Communities; BMI, body mass index; CT, computed tomography; CVD, cardiovascular disease; DXA, dual-energy X-ray absorptiometry; MEC1, Multi-Ethnic Cohort Phase 1; T2DCC, T2D case-control; WC, waist circumference.

At follow-up, a subset of Chinese participants underwent Dual-Energy X-ray Absorptiometry (DXA) and Computed Tomography (CT) scan (9). For the DXA measurements, total body fat mass, trunk fat mass, android fat mass, and lean body mass were assessed using a medium-speed total body acquisition mode (Discovery Wi, Hologic, Bedford, MA, and software Hologic Apex 3.01). For the CT scan, images at the inter-vertebral space L2/L3 level were identified by a single observer (sliceOmatic version 5.0, Tomovision, Magog, Canada). Visceral adipose tissue (VAT) at the L2/L3 level was most strongly correlated with cardiovascular risk factors in Chinese adults (10). The images were analyzed by two independent readers using a workstation (eFilm Workstation version 4.0, Hartland, USA), and readings for VAT area and subcutaneous adipose tissue (SAT) area were averaged. The average inter-observer coefficient of variation was 2.34% for SAT and 3.22% for VAT.

Selected MEC1 participants were profiled on the SomaScan proteomic assay. First, a random sample of 720 participants with equal numbers in each sex and ethnic group were included as the Population Set. The second set included 759 incident type 2 diabetes cases and 1,484 matched controls as the type 2 diabetes case-control (T2DCC) set. Incident type 2 diabetes cases were ascertained through record linkage with a national healthcare database, self-reported physician-diagnosis of diabetes at follow-up, or having a fasting blood glucose ≥ 7mmol/L or random blood glucose ≥ 11mmol/L or A1C ≥ 6.5% at follow-up (11). Controls were selected using risk-set sampling and were matched 1:2 according to age (± 5 years), sex, ethnicity, and date of blood collection (± 2 years). Of 4,897 samples from 2,981 participants sent to SomaLogic, 609 samples were not measured due to shipment issues. We excluded samples that failed SomaLogic quality control, duplicate samples, participants from ethnic groups other than Chinese, Malay, or Indian, participants with a history of heart disease at baseline, and those with missing anthropometric data (**Supplementary Table 1**). The final number of participants for analyses was 631 from the Population Set and 616 cases and 1,200 controls from the T2DCC Set. Written consent was obtained from all participants, and this study was approved by the National University of Singapore Institutional Review Board (reference code: N-18-059).

We externally replicated our findings in the Atherosclerosis Risk in Communities (ARIC) Study, a population-based cohort study in the USA. Participants were male and female adults and white and African American. Details on the ARIC study have been reported elsewhere (12). After the exclusion of participants with diabetes (defined based on self-report diagnosis, medication use, fasting glucose ≥ 7mmol/L, or HbA1c ≥ 6.5%) or heart diseases at baseline, we included a total of 8,428 ARIC participants with proteomic data measured on the SomaScan V4 platform during Visit 2 (1990-1992) in our analyses. Over a median follow-up duration of 19.4 years, 1,800 diabetes incident cases were ascertained based on self-reported diagnosis by a healthcare provider or glucose-lowering medication use reported by participants during annual follow-up telephone calls or clinic visits.

### Proteomic assay

Relative protein abundances were measured in plasma samples using an aptamer-based technology (SomaScan V4 assay) by SomaLogic, Inc. (Boulder, Colorado, US). Details of the SomaScan assay (13,14) and the reproducibility and specificity of the SomaScan assay have been reported previously (15–19).

For this study, relative fluorescence units (RFU) for 5,284 SOMAmers were obtained. After excluding 298 SOMAmers that targeted non-human proteins, 7 deprecated SOMAmers and 1 SOMAmer that targeted a protein whose annotation was withdrawn by the National Center for Biotechnology Information (20), 4,978 SOMAmers targeting 4,775 human proteins remained.

### Statistical analysis

All data analyses were based on log_2_ transformed RFU and winsorized at ±5 standard deviations to reduce the impact of extreme outliers. All P-values reported are from two-sided tests. R version 4.2.0 was used.

### Discovery and internal validation of protein signatures of BMI and WC

Using the Population Set at baseline as the discovery, we evaluated ethnic-specific associations between protein levels and BMI and WC adjusted for age and sex. The ethnic-specific results were pooled using fixed-effects inverse-variance meta-analysis.

We defined associated proteins as those that were (i) significantly associated with the trait of interest (BMI or WC) at the Bonferroni corrected threshold (P < 1.00×10^−05^), (ii) had the same direction of association across three ethnic groups, and (iii) had no evidence of statistical heterogeneity using the Cochran Q test (P_het_ > 1.00×10^−05^). For proteins targeted by multiple SOMAmers, the SOMAmer with the smallest P-value was selected. Using the associated proteins, we performed 100 replicates of 10-fold cross-validation elastic net regression analysis (21). The elastic net regression is a regularized regression method that enables automatic variable selection and variance reduction (21). The BMI-protein and WC-protein signatures were calculated as the weighted sum of the concentration of the selected proteins, with the weights being the beta coefficients from the elastic net regression model.

Using the T2DCC Set as the validation, we examined cross-sectional correlations between the protein signatures and BMI and WC at baseline and follow-up and the correlations between changes in the protein signatures and changes in BMI and WC over a ∼6-year period. On a subset of Chinese participants with DXA measurements (total body fat mass, trunk fat mass, android fat mass, and percentage of lean mass) (n = 207) and VAT and SAT measurements (n = 151), we computed the Pearson correlation coefficients.

### Association between protein signatures and CVD risk factors

We evaluated associations between the protein signatures and systolic blood pressure, HDL cholesterol, LDL cholesterol, triglycerides, fasting glucose, and A1C among participants in the T2DCC Set. We used linear regression models adjusted for age, sex, and ethnicity and standardized all predictor variables to facilitate the comparison of effect sizes. The difference in standardized beta coefficients was evaluated using the Z-test. Participants on anti-hypertensive medication were excluded from analyses involving systolic blood pressure, and participants on lipid-lowering medications were excluded from analyses involving blood lipids.

We evaluated our protein signatures’ ability to differentiate between metabolically healthy and metabolically unhealthy obesity (22) among overweight or obese individuals (BMI ≥ 23.0 kg/m^2^) in the T2DCC set at baseline. We defined metabolically unhealthy as having two or more of the four metabolic abnormalities: (i) hypertension defined as having a systolic blood pressure ≥ 130 mmHg or diastolic blood pressure ≥ 85 mmHg; (ii) elevated fasting plasma glucose, defined as ≥ 5.6 mmol/L, (iii) elevated triglycerides defined as ≥ 1.7 mmol/L, and (iv) reduced HDL cholesterol defined as < 1.0 mmol/L for males and < 1.3 mmol/L for females. We used logistic regression models with metabolic health status as the outcome and standardized protein signatures as predictors, adjusted for age, sex, ethnicity, and BMI.

### Association between protein signatures and type 2 diabetes incidence

We applied the BMI and WC protein signatures on the T2DCC Set to examine the association with type 2 diabetes incidence using logistic regression models adjusted for age, sex, and ethnicity. Variables were standardized to facilitate comparison with BMI and WC.

### External validation of protein signatures in the ARIC study

Using the ARIC study, we first evaluated the associations between anthropometric measurements and each protein in the BMI- and WC-protein signatures, adjusted for age, sex, race, and study center. We consider our adiposity-protein associations replicated if it was statistically significant at the Bonferroni threshold and directionally consistent with the ARIC study. Next, we computed the BMI- and WC-protein signature for each ARIC participant using the weights identified in MEC1 and assessed their correlations with BMI and WC. Finally, we evaluated associations between protein signatures and type 2 diabetes incidence using the Cox proportional hazards model, adjusted for age, sex, race, and study center.

### Functional annotation and pathway enrichment analysis

We performed functional annotation of the proteins using the UniProt Knowledgebase (23). We also queried the Gene Ontology (GO) (24), Kyoto Encyclopedia of Genes and Genomes (KEGG) (25), and Reactome (26) databases using the gprofiler2 R package version 0.2.1 (27) to identify functional annotations and pathways that are significantly overexpressed in the sets of proteins in the BMI- and WC-protein signatures using the hypergeometric test, with all proteins measured by the SomaScan V4 as the custom background. Annotations were considered significant at a false discovery rate (FDR) corrected P-value of 0.05 (28).

### Data and Resource Availability

Summary statistics for all measured proteins are provided in the Supplementary Tables. Researchers can request data from the Multi-Ethnic Cohort study for scientific purposes through an application process at the listed website (https://blog.nus.edu.sg/sphs/data-and-samples-request/). Data will be shared through an institutional data-sharing agreement.

## Results

The baseline characteristics of study participants are shown in **Supplementary Table 2**. Participants were 44.0% male and 56.0% female adults with a mean (± SD) age of 47.6 (± 12.0) years. The ethnic distribution was 34.8% Chinese, 32.6% Malay, and 32.7% Indian.

### Discovery of the BMI and WC-protein signatures

Using the Population Set as discovery, we evaluated the ethnic-specific associations between plasma protein levels with BMI and WC adjusted for age and sex (**Supplementary Table 3**). In the meta-analysis across ethnic groups, 410 proteins were associated with BMI, and 385 proteins were associated with WC, with 359 proteins in common between BMI and WC (**Figure 2**). Across the three ethnic groups, there was no evidence of statistical heterogeneity (P_het_ > 1.00×10^−5^), and all associations were in the same direction except one protein (complement factor D) for WC, which was excluded from further analyses. All 359 overlapping protein-BMI and protein-WC associations were directionally consistent. The top four most strongly associated proteins were the same for BMI and WC, namely leptin (LEP), heart-type fatty acid binding protein (FABP3), insulin-like growth factor-binding protein 1 (IGFBP1), and growth hormone receptor (GHR). We performed a look-up of the 410 BMI-associated proteins in three recent proteomics association studies in European adults, the DIOGenes study (n = 494), the INTERVAL study (n = 2,737), and the KORA study (n = 4,600) (4–6). Out of the 410 BMI-associated proteins observed in our study, 90 proteins were not measured in these three studies, and 263 (82%) out of the remaining 320 BMI-associated proteins were directionally consistent and significant at the Bonferroni threshold in at least one study (**Supplementary Table 3**).

**Figure 2.**
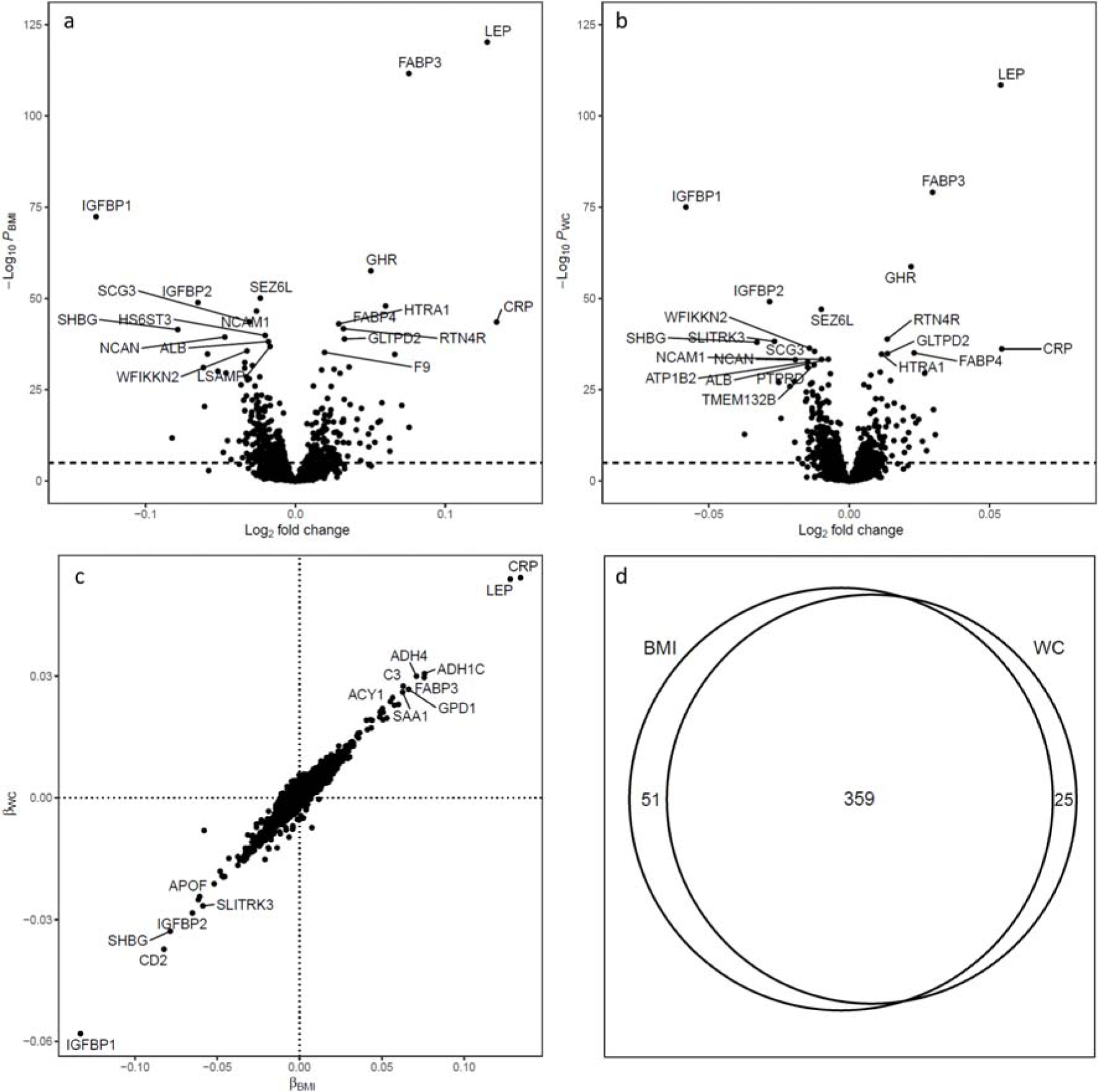
(a) Volcano-plot of −log_10_(P_BMI_) against log_2_(fold change) per kg/m^2^ increment of BMI. (b) Volcano-plot of −log_10_(P_WC_) against log_2_(fold change) per cm increment of WC. Dashed lines represent the threshold for statistical significance after Bonferroni correction. (c) Scatterplot of the beta coefficients for WC against the beta coefficients for BMI. Text annotations are Entrez Gene symbols and the full protein names are provided in Supplementary Table 4. (d) Venn diagram of the number of proteins that were significantly associated with BMI and WC at the Bonferroni threshold. Abbreviations: BMI, body mass index; WC, waist circumference.

We performed feature selection using elastic net regression analysis based on the BMI- and WC-associated proteins. We identified 124 proteins for the BMI-protein signature and 125 proteins for the WC-protein signature, with 60 overlapping proteins in both signatures (**Figure 3** and **Supplementary Table 4**). We compared our protein signatures with the published results of the Fenland Study (15). Of the 124 proteins in our BMI-protein signature, 28 (22.3%) were included in the Fenland Study signature for body fat percentage. Similarly, of the 125 proteins in our WC-protein signature, 34 (27.2%) were included in the Fenland Study signature for visceral fat mass (**Supplementary Table 4)**.

**Figure 3.**
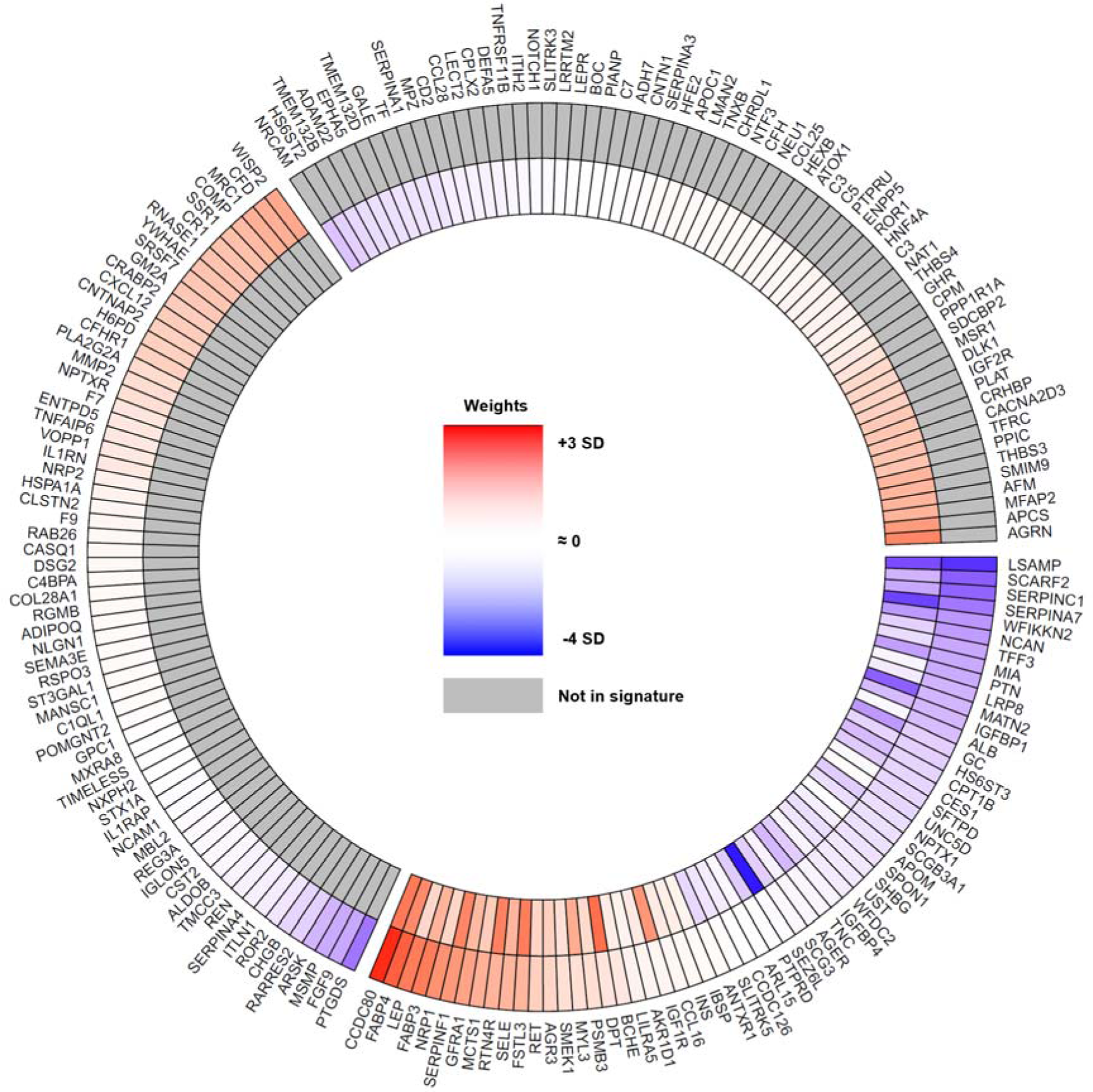
Proteins in the BMI-protein signature (outer circle) and WC-protein signature (inner circle). The colours represent the direction (red = increase, blue = decrease, grey = not in signature) while the intensity of the colours represents the magnitudes of the standardized beta coefficients from elastic net regression. Beta coefficients were standardized to facilitate the comparison of effect sizes across the BMI and WC. The bottom right segment represents the shared proteins between the BMI-protein and WC-protein signature. Moving anticlockwise, the top right segment represents proteins that are only in the WC-protein signature. Finally, the last segment on the top left represents proteins that are only in the BMI-protein signature. Text annotations are Entrez Gene symbols and the full protein names are provided in Supplementary Table 5. Abbreviations: BMI, body mass index; WC, waist circumference.

### Internal validation of the protein signatures

We validated the protein signatures identified in the Population Set using the T2DCC Set. The BMI- and WC-protein signatures were strongly correlated with BMI and WC at baseline and follow-up (r ranging from 0.778 to 0.842) (**Table 1** and **Supplementary Figure 1**). Among 814 participants with both baseline and follow-up data, the changes in BMI (r = 0.632) and WC (r = 0.438) were directly correlated with changes in the respective protein signatures over the same period (**Supplementary Figure 2**). We also examined the correlations between changes in anthropometric measurements and changes in individual proteins in the respective signatures (**Supplementary Table 5**). The Pearson correlation coefficients ranged from 0.438 (leptin) to −0.469 (adiponectin) for changes in BMI and 0.343 (growth hormone receptor) to −0.326 (sex hormone-binding globulin) for changes in WC. All correlation coefficients reported here were significant (P < 0.001).

**Table 1.** Pearson correlation coefficients between body mass index (BMI) and waist circumference (WC) protein signatures and anthropometric, DXA, and CT adiposity measurements in the T2DCC Set.

|  | BMI signature |  |  | WC signature |  |  |
| --- | --- | --- | --- | --- | --- | --- |
|  | Cases | Controls | Combined | Cases | Controls | Combined |
| <u>Anthropometric measurements at baseline *</u> |  |  |  |  |  |  |
| BMI, kg/m<sup>2</sup> | 0.815 | 0.847 | 0.842 | 0.675 | 0.731 | 0.726 |
| WC, cm | 0.639 | 0.717 | 0.716 | 0.675 | 0.798 | 0.778 |
| <u>Anthropometric measurements at follow-up †</u> |  |  |  |  |  |  |
| BMI, kg/m<sup>2</sup> | 0.809 | 0.829 | 0.823 | 0.696 | 0.712 | 0.713 |
| WC, cm | 0.712 | 0.742 | 0.752 | 0.726 | 0.791 | 0.783 |
| <u>DXA measurements ‡</u> |  |  |  |  |  |  |
| Total body fat mass, kg | 0.756 | 0.789 | 0.782 | 0.628 | 0.583 | 0.619 |
| Lean mass, % | -0.482 | -0.484 | -0.488 | -0.270 | -0.171 | -0.232 |
| Android fat mass, kg | 0.755 | 0.787 | 0.787 | 0.706 | 0.689 | 0.717 |
| Trunk fat mass, kg | 0.764 | 0.812 | 0.801 | 0.685 | 0.665 | 0.691 |
| <u>CT measurements §</u> |  |  |  |  |  |  |
| SAT volume, cm<sup>2</sup> | 0.499 | 0.641 | 0.596 | 0.418 | 0.378 | 0.433 |
| VAT volume, cm<sup>2</sup> | 0.542 | 0.719 | 0.676 | 0.603 | 0.776 | 0.735 |

All correlation coefficients were statistically significant at $P < 0.001$ . Abbreviations: BMI, body mass index; Computed Tomography, CT; DXA, Dual-Energy X-ray Absorptiometry; SAT, subcutaneous adipose tissue; T2DCC, type 2 diabetes case control; VAT, visceral adipose tissue; WC, waist circumference.
\* Measured among all participants in the T2DCC Set at baseline; $n = 616$ for cases, 1,200 for controls, and 1,816 when combined.
† Measured among all participants in the T2DCC Set at follow-up; $n = 327$ for cases, 487 for controls, and 814 when combined.
‡ Measured in a subset of Chinese participants at follow-up; $n = 69$ for cases, 138 for controls, and 207 when combined.
§ Measured in a subset of Chinese participants at follow-up; $n = 51$ for cases, 100 for controls, and 151 when combined.

In a subset of Chinese participants with DXA and CT measurements, the BMI-protein signature was directly and strongly correlated with total body fat mass (r = 0.782) and moderately correlated with SAT (r = 0.596) and VAT (r = 0.676) (**Table 1**). In comparison with the BMI-protein signature, the WC-protein signature was less strongly correlated with total body fat mass (r = 0.619) and SAT (r = 0.433) but more strongly correlated with VAT (r = 0.735). The scatterplots suggested a direct linear relationship between total body fat mass and the BMI-protein signature (beta = 4.98 kg per SD; P < 0.001) and VAT area with the WC-protein signature (beta = 48.1 cm^2^ per SD; P < 0.001) (**Figure 4**).

**Figure 4.**
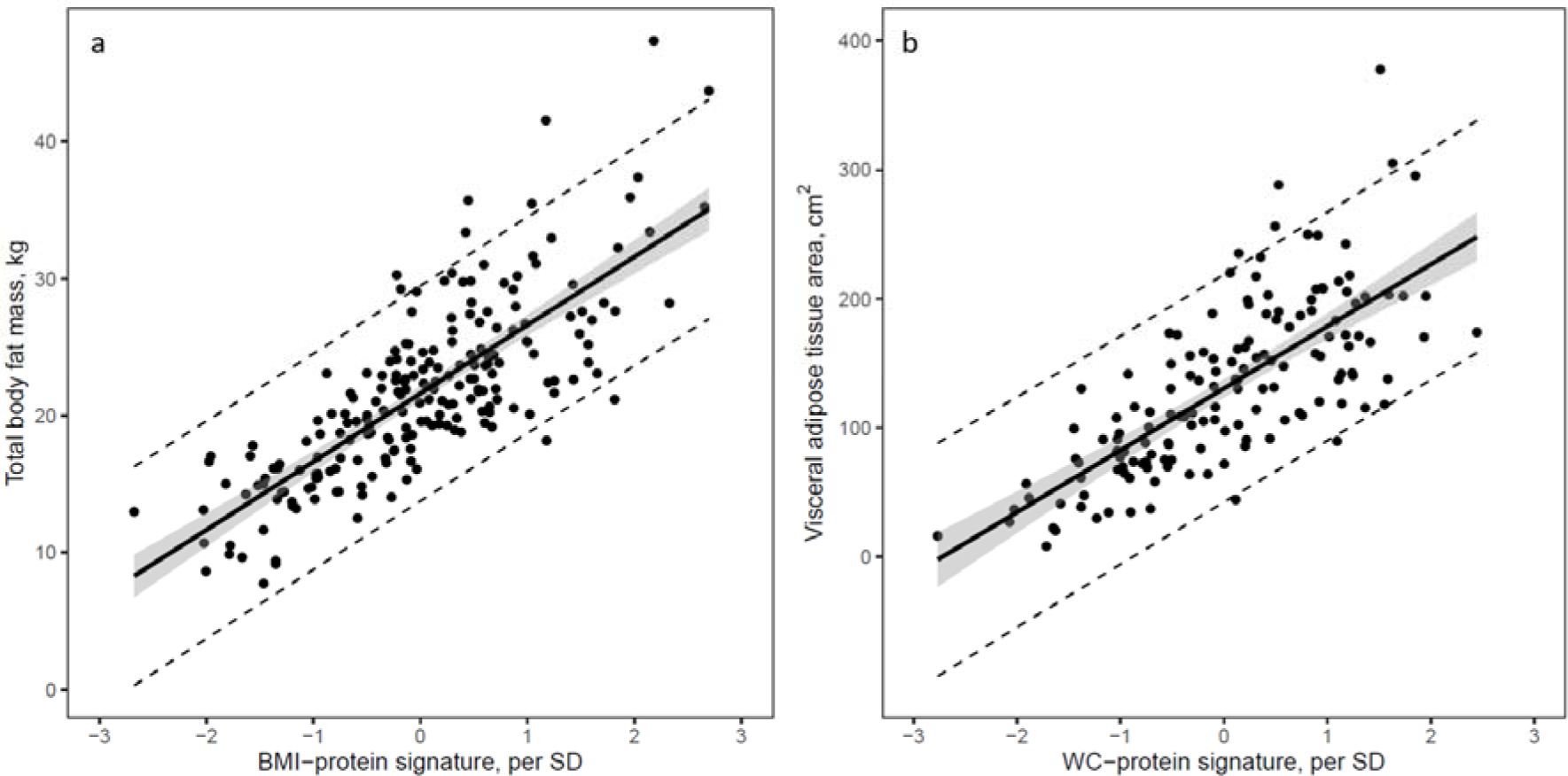
Scatterplots of (a) total body fat mass against BMI-protein signature in 207 Chinese participants with DXA and (b) visceral adipose tissue volume against the WC-protein signature in 151 Chinese participants with CT. Linear regression was fitted with the shaded region representing the 95% confidence intervals and the dotted lines representing the 95% prediction intervals. Abbreviations: CT, computed tomography; DXA, dual-energy X-ray absorptiometry; SD, standard deviation.

### Association between protein signatures and CVD risk factors

In the T2DCC Set, all adiposity-related predictors (BMI, BMI-protein signature, WC, and WC-protein signature) were significantly associated with higher systolic blood pressure, LDL cholesterol, triglycerides, fasting glucose, A1C, and lower HDL cholesterol after adjustment for age, sex, and ethnicity (**Supplementary Table 6**). Protein signatures were significantly more strongly associated with HDL cholesterol, triglycerides, and A1C than anthropometric measurements. Moreover, with the exception of the BMI-protein signature and LDL cholesterol, the associations between the BMI- and WC-protein signatures and CVD risk factors remained statistically significant after further adjustment for BMI or WC (**Supplementary Table 6**).

We also evaluated our protein signatures’ ability to differentiate between metabolically healthy and unhealthy obesity. Among 1,262 overweight (BMI ≥ 23 kg/m^2^) participants in the T2DCC set, 717 (56.8%) were metabolically unhealthy. After adjusting for age, sex, ethnicity, and BMI, having a higher BMI-protein signature (per SD increment, OR = 2.36, 95% CI 1.93 to 2.88) or WC-protein signature (OR = 2.75, 95% CI 2.28 to 3.32) was associated with a higher odds of being metabolically unhealthy. Results were similar when we used the international BMI cut-off of 25.0 kg/m^2^ to define overweight.

### Association between protein signatures and type 2 diabetes incidence

In the T2DCC Set, the BMI-protein signature (OR 2.44, 95% CI 2.15 to 2.77) and WC-protein signature (OR 2.84, 95% CI 2.47 to 3.25) were significantly associated with type 2 diabetes incidence, with larger effect sizes (P_diff_ < 0.05) than BMI (OR 1.91, 95% CI 1.70 to 2.14) and WC (OR 2.09, 95% CI 1.86 to 2.36) (**Table 2**). Moreover, the associations between the BMI- and WC-protein signatures and type 2 diabetes remained significant after further adjustment for BMI and WC, respectively.

**Table 2.** Associations between anthropometric measurements, protein signatures of adiposity, and the incidence of type 2 diabetes.

|  | **OR (95% CI)** | **P-value** | **P<sub>diff</sub> \*** |
| --- | --- | --- | --- |
| **BMI** | 1.91 (1.70, 2.14) | 1.95E-28 | 0.005 |
| **BMI-protein signature** | 2.44 (2.15, 2.77) | 1.19E-42 |  |
| **BMI-protein signature adjusted for BMI** | 2.69 (2.18, 3.32) | 3.80E-20 |  |
| **WC** | 2.09 (1.86, 2.36) | 2.09E-33 | 0.001 |
| **WC-protein signature** | 2.84 (2.47, 3.25) | 1.64E-50 |  |
| **WC-protein signature adjusted for WC** | 2.69 (2.23, 3.25) | 6.25E-25 |  |

All predictor variables were expressed as per 1 SD increment to facilitate the comparison of effect sizes. All models were adjusted for age, sex, and ethnicity.
Abbreviations: BMI, body mass index; WC, waist circumference.
\* P-value for difference between the standardized coefficients for BMI and the BMI-protein signature, and the coefficients between WC and the WC-protein signature, calculated using the Z test.

As the effects of adiposity on insulin resistance may be modified by ethnicity (29,30), we conducted further analyses stratified by ethnicity (**Supplementary Table 7**). We found a stronger association between the BMI-protein signature and type 2 diabetes among Malays (OR 2.96, 95% CI 2.33 to 3.77) and Chinese (OR 2.37, 95% CI 1.94 to 2.88) than Indians (OR 1.91, 95% CI 1.55 to 2.35) (P-interaction = 0.019). Similarly, the WC-protein signature was more strongly associated with type 2 diabetes incidence among Malays (OR 3.59, 95% CI 2.77 to 4.67) and Chinese (OR 2.72, 95% CI 2.20 to 3.37) than Indians (OR 2.21, 95% CI 1.76 to 2.77) (P-interaction = 0.028).

### External validation in the ARIC study

Participants in the ARIC study had a mean age of 56.7 (± 5.7) years, were mostly female (57%), and were white (81%) or African African (19%) (**Supplementary Table 8**). After Bonferroni adjustment (P < 0.05/124 for BMI and 0.05/125 for WC), 118 (95.2%) of 124 proteins in the BMI-protein signature and 124 (99.2%) of 125 proteins in the WC-protein signature were directionally consistent and significantly associated with BMI and WC, respectively (**Supplementary Table 4** and **Supplementary Figure 3**). Concordant with findings in MEC1, the BMI-protein signature was highly correlated with BMI (r = 0.822), and the WC-protein signature was highly correlated with WC (r = 0.773). A higher BMI-protein signature (HR per SD increment = 1.80, 95% CI 1.72 to 1.89) and WC-protein signature (HR = 1.97, 95% CI 1.87 to 2.07) at baseline were strongly associated with type 2 diabetes incidence over a median follow-up of 19.4 years.

### Pathway enrichment analysis

In our pathway enrichment analysis, we identified 36 significant annotations from the BMI-protein signature and 28 significant annotations from the WC-protein signature (**Supplementary Table 9**). Pathways related to post-translational protein phosphorylation, the regulation of insulin-like growth factor (IGF) transport and uptake by insulin-like growth factor binding proteins (IGFBPs), and complement and coagulation cascades were significantly enriched (P_FDR_ < 0.05) in both BMI- and WC-protein signatures. In additional, pathways involved in adenosine monophosphate-activated protein kinase (AMPK) signaling, extracellular matrix (ECM) receptor interaction, and cell adhesion molecules (CAMs)– were significantly enriched (P_FDR_ < 0.05) in the WC-protein signature.

## Discussion

We identified 410 proteins associated with BMI and 385 proteins associated with waist circumference that were consistent across all three Asian ethnic groups (Chinese, Malay, and Indian) in our study population. We derived protein signatures of BMI and WC which we validated in an independent dataset across two timepoints and externally in a cohort of US white and African American participants. Compared to anthropometric measurements, our protein signatures were more strongly associated with cardiometabolic risk factors and were able to distinguish between metabolically healthy and unhealthy obesity. In prospective analyses, both protein signatures were strongly associated with the risk of type 2 diabetes in MEC1 and the ARIC study. Biological pathways related to post-translational protein phosphorylation, regulation of IGFs and IGFBPs, coagulation cascades, AMPK signaling, ECM receptor interaction, and cell adhesion were overrepresented in the BMI- and WC-protein signatures.

Of the 410 proteins associated with BMI in our Asian cohort, over 80% of the proteins measured in common were replicated in previous studies in European populations (4–6), highlighting the robustness of these findings to ethnic and geographical variation. In addition, we identified novel proteins that were not previously reported, most likely because of the larger number of proteins represented in the proteomic assay used in our study. These include serine protease high temperature requirement A1 (HTRA1), syndecan-3 (SDC3), and complexin-2 (CPLX2). HTRA1 has been shown to regulate the availability of IGFs (31,32). In addition, HTRA1 may inhibit adipogenesis by regulating the formation of adipocytes and could be an indicator of adipose tissue dysfunction (33). Variants of the SDC3 gene have been associated with obesity in Korean (34) and Taiwanese adults (35). SDC3 is involved in regulating appetite through the melanocortin system (36) and may, therefore, be a potential therapeutic target for treating obesity. CPLX2 may affect the translocation of glucose transporter 4 (GLUT4), which plays a critical role in glucose uptake regulation (37). However, it should be noted that GLUT4 translocation is an intracellular process, and whether plasma levels of CPLX2 are related to this process is unclear. In addition, CPLX2 may also have a role in the regulation of the immune system (38). These novel adiposity-related proteins, if replicated, can inform research on the pathophysiology of obesity and related cardiometabolic conditions.

Using supervised machine learning techniques, we identified protein signatures of BMI and WC and validated these across two timepoints in an independent dataset and externally in the ARIC study. A large proportion (85%) of the proteins significantly associated with WC and BMI in MEC1 overlapped. However, greater differences existed in proteins selected by machine learning for the BMI and WC signatures, with slightly less than half of the proteins overlapping. We further showed that the BMI-protein signature was highly correlated with total body fat mass, whereas the WC-protein signature was more strongly correlated with VAT. These findings suggest that the two protein signatures were able to capture the differences in plasma proteins associated with adiposity in different anatomical compartments.

Compared to anthropometric measures, the BMI- and WC-protein signatures were more strongly associated with HDL-cholesterol, triglycerides, hemoglobin A1c levels, and the incidence of type 2 diabetes. In addition, the associations of these protein signatures with CVD risk factors and type 2 diabetes remained significant after further adjustment for BMI or WC. These findings suggest that the protein signatures are likely to capture physiologically relevant effects of body fat better than traditional measures of adiposity.

The identified plasma protein signatures differentiated between metabolically healthy and metabolically unhealthy obesity and were associated with type 2 diabetes risk. Thus, the biological pathways represented by the adiposity-related proteins may provide mechanistic insights in the physiological changes associated with metabolically unhealthy obesity (39). Pathways related to complement and coagulation cascades, IGFs and IGFBPs regulation, and post-translational protein phosphorylation were significantly enriched in both the BMI-and WC-protein signatures. In addition, pathways involving AMPK signaling, ECM-receptor interaction, and CAMs were enriched in the WC-protein signature. The dysregulation of the complement system resulting from overexpression of cytokines and adipose tissue-derived factors may lead to chronic inflammation and the development of metabolic disorders such as insulin resistance and type 2 diabetes (40). Similarly, imbalances in levels of IGFs and IGFBPs have been linked to the dysregulation of fat metabolism and insulin resistance (41–43). AMPK is a key enzyme involved in regulating energy and nutrient metabolism, and the dysregulation of AMPK has been implicated in the development of obesity, insulin resistance, and type 2 diabetes (44). The over-activation of ECM receptor signaling pathways may lead to inflammation and insulin resistance (45). Elevation of CAMs has been previously associated with obesity and type 2 diabetes (46,47) and may affect inflammation and atherogenesis (48). Taken together, the pathways enriched in the adiposity-related protein signatures depict a state of dysregulated cell signaling, systemic inflammation, and impaired glucose and fat metabolism. In addition, several of the pathways (e.g., complement pathways, ECM-receptor interaction, and IGFBP regulation) enriched in our adiposity-related protein signatures were also enriched in type 2 diabetes-associated proteins in European populations (49), underscoring the central role of obesity-related pathways in the pathogenesis of type 2 diabetes.

Previous studies have reported ethnic differences in the association between obesity and insulin resistance (29,50). Khoo et al. (29) demonstrated that the association of body fat percentage with insulin resistance was weaker in ethnic Indians than in Chinese and Malays residing in Singapore. Furthermore, Retnakaran et al. (50) found that pre-pregnancy BMI was more strongly associated with insulin resistance in East Asian women than in South Asian women in Canada. Here, while associations between plasma proteins and adiposity measures were consistent across ethnic groups, we observed stronger associations between the BMI- and WC-protein signatures and type 2 diabetes risk in Chinese and Malays than Indians.

These findings suggest that ethnic differences in the associations between obesity and type 2 diabetes may be explained by differential effects of obesity-related proteins on type 2 diabetes.

We acknowledge the limitations of our study. First, the plasma proteome including both secreted and leakage proteins has been shown to be informative about the current and future state of health (15,19), but the concentration of leakage proteins in the plasma may not be a direct reflection of its biological activity in the cells. Second, we were not able to infer causality as the associations between plasma proteins and anthropometric measurements were cross-sectional. Even though our participants did not have type 2 diabetes at baseline, early manifestations of diabetes may have influenced plasma protein levels. Third, while studies have demonstrated the specificity of the SomaScan assay (18,19), it remains possible that some proteins may not be correctly identified by the SomaScan platform used in our study.

In conclusion, our findings on adiposity-related proteins were robust across ethnic and geographically diverse groups and have potential uses for assessing and monitoring metabolically adverse adiposity. While further studies are needed to evaluate the causal relationship between the identified proteins and adiposity, the results from this study can lead to a better understanding of the biological mechanisms relating obesity and type 2 diabetes and why sub-groups of the population differ in their susceptibility to adverse effects of obesity on metabolic health.

## Data Availability

Summary statistics for all measured proteins are provided in the Supplementary Tables. Data from the Multi-Ethnic Cohort study can be requested by researchers for scientific purposes through an application process at the listed website (https://blog.nus.edu.sg/sphs/data-and-samples-request/). Data will be shared through an institutional data sharing agreement.

## Acknowledgments

We thank all participants, the study team, and the investigators for their research contributions. The MEC1 study is supported by individual research and clinical scientist award schemes from the Singapore National Medical Research Council (NMRC, including MOH-000271-00) and the Singapore Biomedical Research Council (BMRC), the Singapore Ministry of Health (MOH), the National University of Singapore (NUS) and the Singapore National University Health System (NUHS). The Atherosclerosis Risk in Communities study has been funded in whole or in part with Federal funds from the National Heart, Lung, and Blood Institute, National Institutes of Health, Department of Health and Human Services, under Contract nos. (75N92022D00001, 75N92022D00002, 75N92022D00003, 75N92022D00004, 75N92022D00005). The funders had no role in the design, implementation, analysis, and interpretation of the data.

## Author contributions

X.S., and R.M.vD designed the study. C.G.Y.L., X.S., R.M.vD, B.O., M.R.R., C.E.N., J.C., J.Y., Y.L., E.S.T., N.E.E.K. contributed to the acquisition, analysis, or interpretation of data. C.G.Y.L., X.S., R.M.vD. contributed to the drafting of manuscript. E.S.T., B.O., M.R.R., C.E.N., J.C. contributed to the critical revision of manuscript. X.S. is the guarantor of this work and, as such, had full access to all the data in the study and takes responsibility for the integrity of the data and the accuracy of the data analysis.

## Disclaimers

The National University of Singapore has signed a collaboration agreement with SomaLogic to conduct SomaScan of MEC stored samples at no charge in exchange for the rights to analyze linked MEC phenotype data.

## Supplementary Figures

**Supplementary Figure 1.**
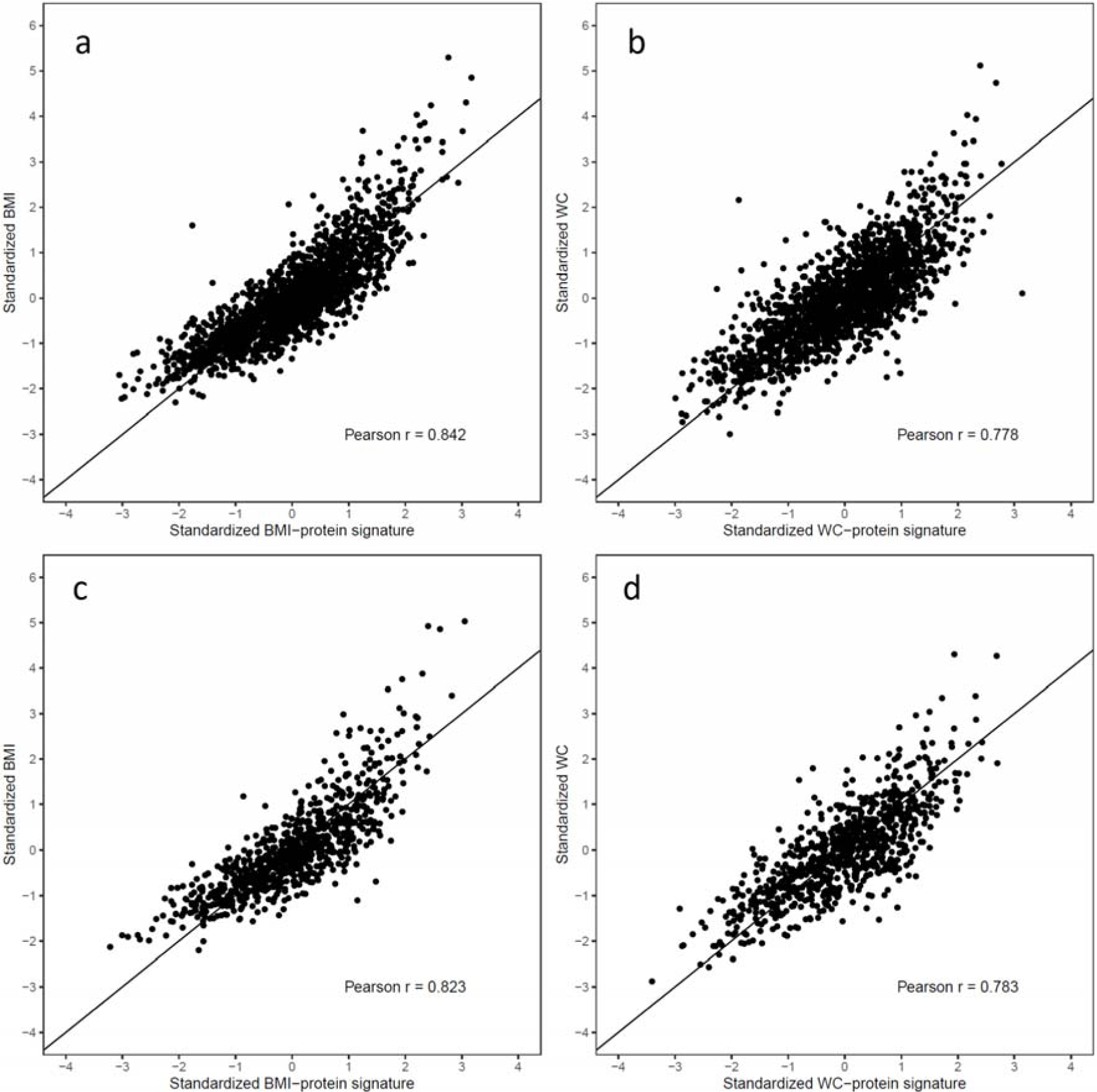
Scatterplots of (a) BMI against BMI-protein signature at baseline (b) WC against WC-protein signature at baseline, (c) BMI against BMI-protein signature at follow-up, and (d) WC against WC-protein signature at follow-up in the T2DCC Set. The diagonal lines represent the y = x line. Abbreviations: BMI, body mass index; T2DCC, type 2 diabetes case control; WC, waist circumference.

**Supplementary Figure 2.**
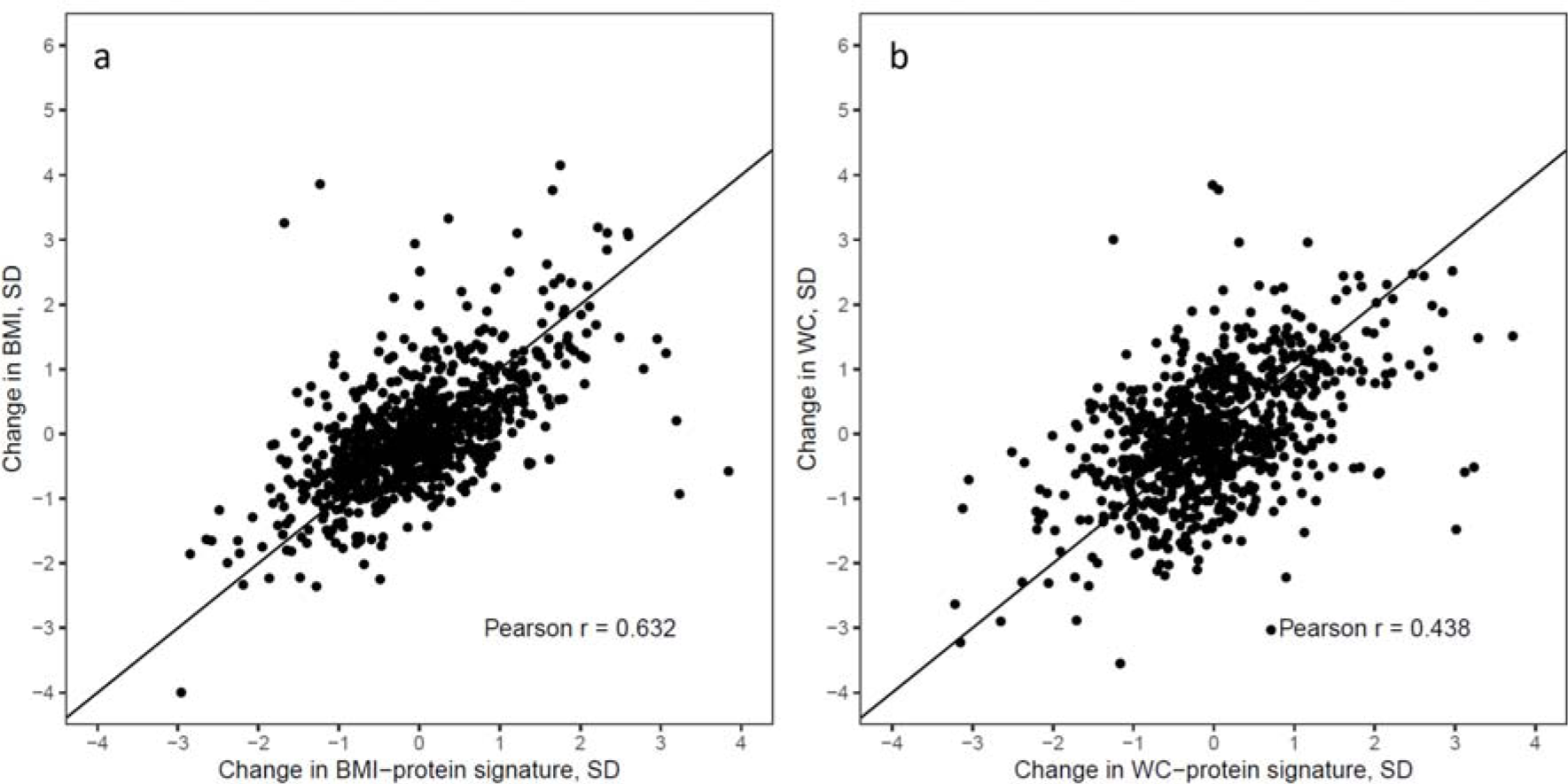
Scatterplots of (a) change in BMI against change in BMI-protein signature, and (b) change in WC against change in WC-protein signature over a ∼6-year period in the T2DCC set. The diagonal lines represent the y = x line. Abbreviations: BMI, body mass index; SD, standard deviation; T2DCC, type 2 diabetes case control; WC, waist circumference.

**Supplementary Figure 3.**
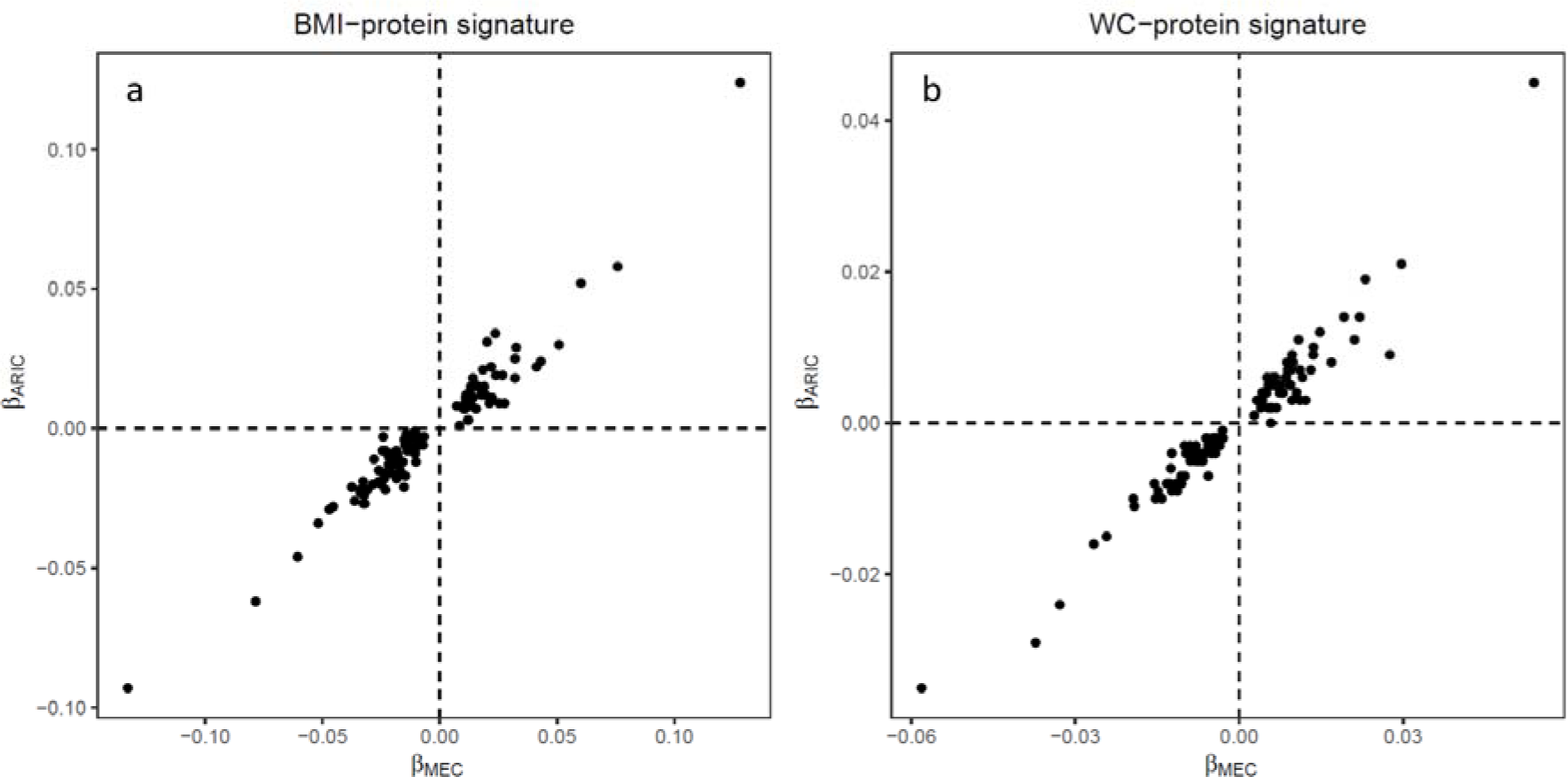
Comparison of the beta coefficients for the associations between plasma proteins and BMI/WC between the Singapore Multi-ethnic cohort Phase 1 (MEC1) and the Atherosclerosis Risk in Communities (ARIC) study for (a) the BMI-protein signature and (b) the WC-protein signature. Plasma protein levels in relative fluorescence units (RFU) were quantified using the SomaScan V4 assay in both studies. Each point corresponds to each protein in the respective protein signatures (n = 124 for the BMI-protein signature and n = 125 for the WC-protein signature). Abbreviations: BMI, body mass index; WC, waist circumference.

## References

1. Hruby A, Hu FB. The Epidemiology of Obesity: A Big Picture. Pharmacoeconomics. 2015 Dec 4;33(7):673–89.

2. Riaz H, Khan MS, Siddiqi TJ, Usman MS, Shah N, Goyal A, et al. Association Between Obesity and Cardiovascular Outcomes: A Systematic Review and Meta-analysis of Mendelian Randomization Studies. JAMA Netw Open. 2018 Nov 2;1(7):e183788–e183788.

3. Hanash S. Disease proteomics. Nature. 2003 Mar 13;422(6928):226–32.

4. Carayol J, Chabert C, Di Cara A, Armenise C, Lefebvre G, Langin D, et al. Protein quantitative trait locus study in obesity during weight-loss identifies a leptin regulator. Nat Commun. 2017 Dec 1;8(1).

5. Zaghlool SB, Sharma S, Molnar M, Matías-García PR, Elhadad MA, Waldenberger M, et al. Revealing the role of the human blood plasma proteome in obesity using genetic drivers. Nat Commun. 2021 Feb 24;12(1):1–13.

6. Goudswaard LJ, Bell JA, Hughes DA, Corbin LJ, Walter K, Davey Smith G, et al. Effects of adiposity on the human plasma proteome: observational and Mendelian randomisation estimates. Int J Obes (Lond). 2021 Oct;45(10):2221–9.

7. Ma RCW, Chan JCN. Type 2 diabetes in East Asians: similarities and differences with populations in Europe and the United States. Ann N Y Acad Sci. 2013;1281(1):64–91.

8. Tan KHX, Tan LWL, Sim X, Tai ES, Lee JJM, Chia KS, et al. Cohort Profile: The Singapore Multi-Ethnic Cohort (MEC) study. Int J Epidemiol. 2018;47(3):699–699j.

9. Khaing NEE, Shyong TE, Lee J, Soekojo CY, Ng A, Van Dam RM. Epicardial and visceral adipose tissue in relation to subclinical atherosclerosis in a Chinese population. PLoS One. 2018 Apr 1;13(4):e0196328.

10. Ng AC, Wai DC, Tai ES, Ng KM, Chan LL. Visceral adipose tissue, but not waist circumference is a better measure of metabolic risk in Singaporean Chinese and Indian men. Nutr Diabetes. 2012 Aug 6;2(8):e38–e38.

11. American Diabetes Association. 2. Classification and Diagnosis of Diabetes: Standards of Medical Care in Diabetes—2021. Diabetes Care. 2021 Jan 1;44(Supplement_1):S15–33.

12. Steffen BT, Tang W, Lutsey PL, Demmer RT, Selvin E, Matsushita K, et al. Proteomic analysis of diabetes genetic risk scores identifies complement C2 and neuropilin-2 as predictors of type 2 diabetes: the Atherosclerosis Risk in Communities (ARIC) Study. Diabetologia. 2022 Jan 1;66(1):105–15.

13. Rohloff JC, Gelinas AD, Jarvis TC, Ochsner UA, Schneider DJ, Gold L, et al. Nucleic Acid Ligands With Protein-like Side Chains: Modified Aptamers and Their Use as Diagnostic and Therapeutic Agents. Mol Ther Nucleic Acids. 2014;3(10):e201.

14. Gold L, Ayers D, Bertino J, Bock C, Bock A, Brody EN, et al. Aptamer-Based Multiplexed Proteomic Technology for Biomarker Discovery. PLoS One. 2010;5(12):e15004.

15. Williams SA, Kivimaki M, Langenberg C, Hingorani AD, Casas JP, Bouchard C, et al. Plasma protein patterns as comprehensive indicators of health. Nat Med. 2019 Dec 2;25(12):1851–7.

16. Kim CH, Tworoger SS, Stampfer MJ, Dillon ST, Gu X, Sawyer SJ, et al. Stability and reproducibility of proteomic profiles measured with an aptamer-based platform. Sci Reports 2018 81. 2018 May 30;8(1):1–10.

17. Tin A, Yu B, Ma J, Masushita K, Daya N, Hoogeveen RC, et al. Reproducibility and Variability of Protein Analytes Measured Using a Multiplexed Modified Aptamer Assay. J Appl Lab Med. 2019 Jul 1;4(1):30.

18. Sun BB, Maranville JC, Peters JE, Stacey D, Staley JR, Blackshaw J, et al. Genomic atlas of the human plasma proteome. Nature. 2018 Jun 6;558(7708):73–9.

19. Emilsson V, Ilkov M, Lamb JR, Finkel N, Gudmundsson EF, Pitts R, et al. Co-regulatory networks of human serum proteins link genetics to disease. Science. 2018 Aug 8;361(6404):769.

20. NCBI. LOC652493 ig kappa chain V-I region HK102-like [Homo sapiens (human)] - Gene - NCBI [Internet]. 2011.

21. Zou H, Hastie T. Regularization and variable selection via the elastic net. J R Stat Soc Ser B (Statistical Methodol. 2005 Apr 1;67(2):301–20.

22. Blüher M. Metabolically Healthy Obesity. Endocr Rev. 2020 Jun 1;41(3):405–20.

23. Bateman A, Martin MJ, Orchard S, Magrane M, Ahmad S, Alpi E, et al. UniProt: the Universal Protein Knowledgebase in 2023. Nucleic Acids Res. 2023 Jan 6;51(D1):D523–31.

24. Ashburner M, Ball CA, Blake JA, Botstein D, Butler H, Cherry JM, et al. Gene Ontology: tool for the unification of biology. Nat Genet. 2000 May;25(1):25.

25. Kanehisa M, Goto S. KEGG: Kyoto Encyclopedia of Genes and Genomes. Nucleic Acids Res. 2000 Jan 1;28(1):27–30.

26. Fabregat A, Jupe S, Matthews L, Sidiropoulos K, Gillespie M, Garapati P, et al. The Reactome Pathway Knowledgebase. Nucleic Acids Res. 2018 Jan 1;46(Database issue):D649.

27. Kolberg L, Raudvere U, Kuzmin I, Vilo J, Peterson H. gprofiler2 -- an R package for gene list functional enrichment analysis and namespace conversion toolset g:Profiler. F1000Research. 2020 Jul 15;9:709.

28. Benjamini Y, Hochberg Y. Controlling the False Discovery Rate: A Practical and Powerful Approach to Multiple Testing. J R Stat Soc Ser B. 1995 Jan 1;57(1):289–300.

29. Khoo CM, Khee-Shing Leow M, Sadananthan SA, Lim R, Venkataraman K, Khoo EYH, et al. Body Fat Partitioning Does Not Explain the Interethnic Variation in Insulin Sensitivity Among Asian Ethnicity: The Singapore Adults Metabolism Study. Diabetes. 2014 Mar 1;63(3):1093–102.

30. Khoo CM, Sairazi S, Taslim S, Gardner D, Wu Y, Lee J, et al. Ethnicity Modifies the Relationships of Insulin Resistance, Inflammation, and Adiponectin With Obesity in a Multiethnic Asian Population. Diabetes Care. 2011 May 1;34(5):1120–6.

31. Hou J, Clemmons DR, Smeekens S. Expression and characterization of a serine protease that preferentially cleaves insulin-like growth factor binding protein-5. J Cell Biochem. 2005 Feb 15;94(3):470–84.

32. Jacobo SMP, DeAngelis MM, Kim IK, Kazlauskas A. Age-Related Macular Degeneration-Associated Silent Polymorphisms in HtrA1 Impair Its Ability To Antagonize Insulin-Like Growth Factor 1. Mol Cell Biol. 2013 May 1;33(10):1976.

33. Tiaden AN, Bahrenberg G, Mirsaidi A, Glanz S, Blüher M, Richards PJ. Novel Function of Serine Protease HTRA1 in Inhibiting Adipogenic Differentiation of Human Mesenchymal Stem Cells via MAP Kinase-Mediated MMP Upregulation. Stem Cells. 2016 Jun 1;34(6):1601–14.

34. Ha E, Kim MJ, Choi BK, Rho JJ, Oh DJ, Rho TH, et al. Positive Association of Obesity with Single Nucleotide Polymorphisms of Syndecan 3 in the Korean Population. J Clin Endocrinol Metab. 2006 Dec 1;91(12):5095–9.

35. Chang BCC, Hwang LC, Huang WH. Positive Association of Metabolic Syndrome with a Single Nucleotide Polymorphism of Syndecan-3 (rs2282440) in the Taiwanese Population. Int J Endocrinol. 2018;2018.

36. Reizes O, Benoit SC, Strader AD, Clegg DJ, Akunuru S, Seeley RJ. Syndecan-3 Modulates Food Intake by Interacting with the Melanocortin/AgRP Pathway. Ann N Y Acad Sci. 2003 Jun 1;994(1):66–73.

37. Pavarotti MA, Tokarz V, Frendo-Cumbo S, Bilan PJ, Liu Z, Zanni-Ruiz E, et al. Complexin-2 redistributes to the membrane of muscle cells in response to insulin and contributes to GLUT4 translocation. Biochem J. 2021 Jan 29;478(2):407–22.

38. Tsuru E, Oryu K, Sawada K, Nishihara M, Tsuda M. Complexin 2 regulates secretion of immunoglobulin in antibody-secreting cells. Immunity, Inflamm Dis. 2019 Dec 1;7(4):318–25.

39. Iacobini C, Pugliese G, Blasetti Fantauzzi C, Federici M, Menini S. Metabolically healthy versus metabolically unhealthy obesity. Metabolism. 2019 Mar 1;92:51–60.

40. Shim K, Begum R, Yang C, Wang H. Complement activation in obesity, insulin resistance, and type 2 diabetes mellitus. World J Diabetes. 2020 Jan 1;11(1):1.

41. Friedrich N, Thuesen B, Jrøgensen T, Juul A, Spielhagen C, Wallaschofksi H, et al. The Association Between IGF-I and Insulin Resistance: A general population study in Danish adults. Diabetes Care. 2012 Apr;35(4):768.

42. Haywood NJ, Slater TA, Matthews CJ, Wheatcroft SB. The insulin like growth factor and binding protein family: Novel therapeutic targets in obesity & diabetes. Mol Metab. 2019 Jan 1;19:86.

43. Clemmons DR. Role of IGF-binding proteins in regulating IGF responses to changes in metabolism. J Mol Endocrinol. 2018 Jul 1;61(1):T139–69.

44. Steinberg GR, Hardie DG. New insights into activation and function of the AMPK. Nat Rev Mol Cell Biol. 2023 Apr;24(4):255–72.

45. Lin D, Chun TH, Kang L. Adipose extracellular matrix remodelling in obesity and insulin resistance. Biochem Pharmacol. 2016 Nov 1;119:8–16.

46. Hegazy GA, Awan Z, Hashem E, Al-Ama N, Abunaji AB. Levels of soluble cell adhesion molecules in type 2 diabetes mellitus patients with macrovascular complications. J Int Med Res. 2020;48(4):1–11.

47. Miller MA, Cappuccio FP. Cellular adhesion molecules and their relationship with measures of obesity and metabolic syndrome in a multiethnic population. Int J Obes 2006 308. 2006 Mar 7;30(8):1176–82.

48. Hope SA, Meredith IT. Cellular adhesion molecules and cardiovascular disease. Part I. Their expression and role in atherogenesis. Intern Med J. 2003 Aug 1;33(8):380–6.

49. Gudmundsdottir V, Zaghlool SB, Emilsson V, Aspelund T, Ilkov M, Gudmundsson EF, et al. Circulating Protein Signatures and Causal Candidates for Type 2 Diabetes. Diabetes. 2020 Aug 1;69(8):1843.

50. Retnakaran R, Hanley AJG, Connelly PW, Sermer M, Zinman B. Ethnicity Modifies the Effect of Obesity on Insulin Resistance in Pregnancy: A Comparison of Asian, South Asian, and Caucasian Women. J Clin Endocrinol Metab. 2006 Jan 1;91(1):93–7.

